## Supplementary figures for "Reverse mutational scanning of spike BA.2.86 identifies epitopes contributing to immune escape from polyclonal sera"

### Supplementary figure 1

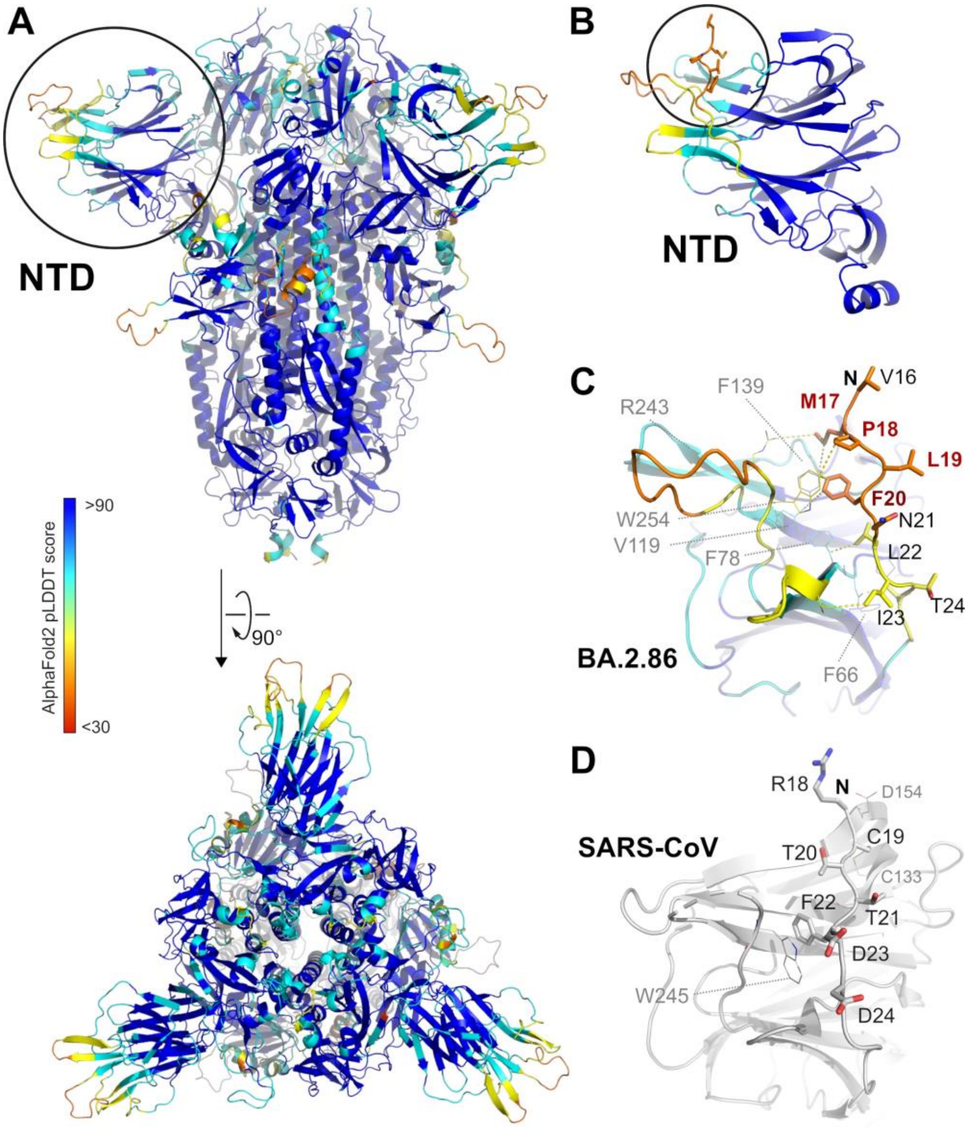


**Figure S1. Analysis of the structural model of the BA.2.86 spike protein**

(A) AlphaFold2/AlphaFold-Multimer model of the trimeric spike protein, colored by per-residue model confidence score (pLDDT), with blue colors corresponding to high and red colors corresponding to low confidence. The circle highlights the N-terminal domain NTD further explored in the next panel. (B) The NTD of BA.2.86 spike protein colored by pLDDT score. The circle highlights the N-terminal 16MPLF insertion of BA.2.86 and is further explored in the next panel. (C) Predicted interactions of the 16MPLF insertion with a crevice of the NTD. Colors correspond to the pLDDT score, N marks the position of the N-terminus after cleavage of the signal peptide. (D) Comparison to the same region in the NTD of SARS-CoV (PDB-ID 5X4S) (28). Note that this virus, with respect to SARS-CoV-2, carried an extended N-terminus as well. Here, the N-terminus was anchored by a disulfide bridge (C19-C133), which is not conserved in BA.2.86.

### Supplementary figure 2

#### A) Neutralisation titers for NTD specific BA.2.86pp mutants post XBB.1.5 adapted vaccine

| | | | | | | | | | | | | | | | |
|---|---|---|---|---|---|---|---|---|---|---|---|---|---|---|---|
| Geometric mean titer: | 7969 | 1280 | 4986 | 1074 | 588 | 933 | 802 | 1801 | 1843 | 941 | 859 | 972 | 1492 | 1157 | 1487 |
| Fold reduction (vs. BA.2): | - | 6 | 2 | 7 | 14 | 9 | 10 | 4 | 4 | 8 | 9 | 8 | 5 | 7 | 5 |
| Sample number: | 30 | 30 | 24 | 28 | 28 | 24 | 25 | 25 | 21 | 28 | 28 | 25 | 26 | 27 | 28 |

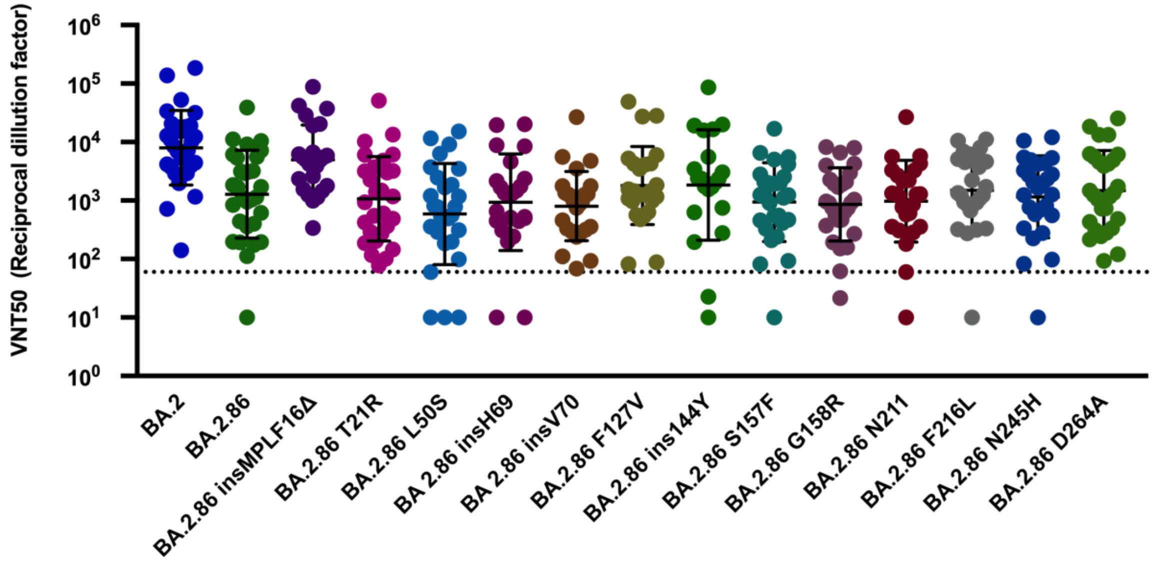


#### B) Neutralisation titers for RBD specific BA.2.86pp mutants post XBB.1.5 adapted vaccine

| | | | | | | | | | | | | | | | | |
|---|---|---|---|---|---|---|---|---|---|---|---|---|---|---|---|---|
| Geometric mean titer: | 7969 | 1280 | 625 | 1020 | 1869 | 401 | 1072 | 1829 | 854 | 833 | 3197 | 1831 | 2182 | 4196 | 2060 | 289 |
| Fold reduction (vs. BA.2): | - | 6 | 13 | 8 | 4 | 20 | 7 | 4 | 9 | 10 | 2 | 4 | 4 | 2 | 4 | 28 |
| Sample number: | 30 | 30 | 27 | 24 | 25 | 28 | 28 | 20 | 24 | 24 | 28 | 24 | 20 | 20 | 27 | 24 |

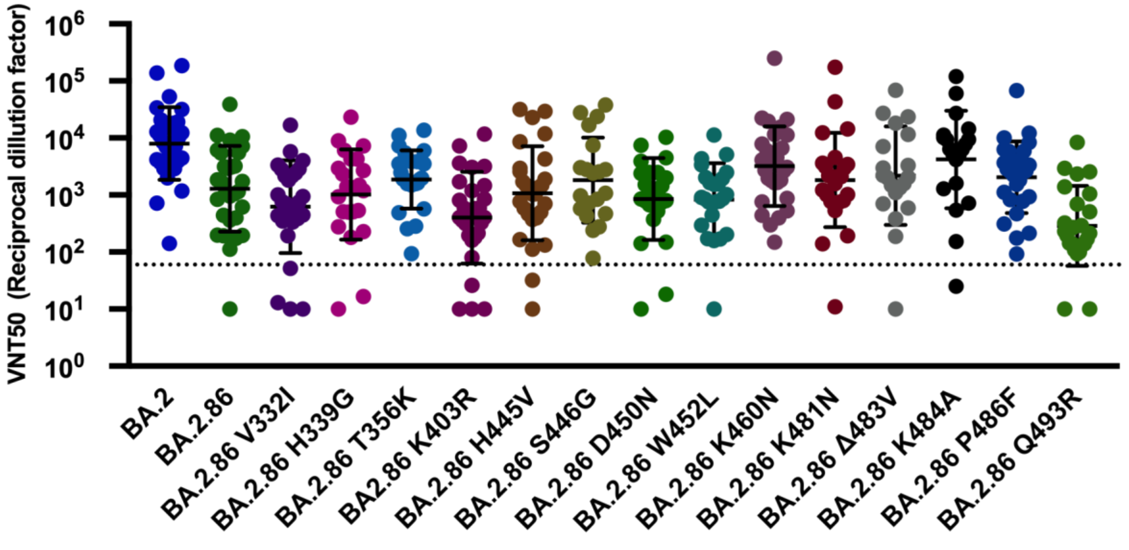

### Supplementary figure 2

**C)** **Neutralisation titers for S1/S2 specific BA.2.86pp mutants post XBB.1.5 adapted vaccine**

| | | | | | | | | |
|---|---|---|---|---|---|---|---|---|
| Geometric mean titer: | 7969 | 1280 | 2706 | 1014 | 2834 | 1040 | 1844 | 830 |
| Fold reduction (vs. BA.2): | - | 6 | 3 | 8 | 3 | 8 | 4 | 10 |
| Sample number: | 30 | 30 | 24 | 28 | 24 | 17 | 20 | 24 |

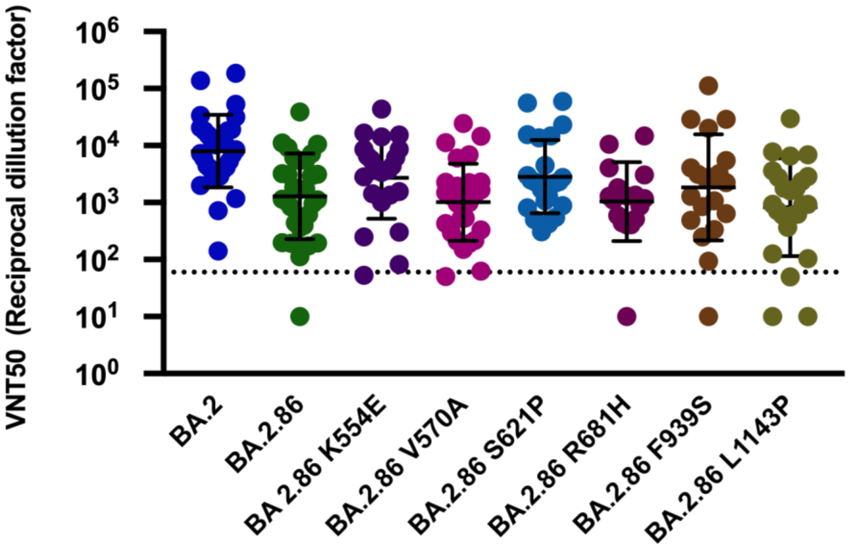


**Figure S2. Neutralization titer of plasma samples post vaccination with the BNT162b2 XBB.1.5 adapted vaccine.**

Neutralization titer of plasma samples post vaccination with the BNT162b2 XBB.1.5 adapted vaccine against (A) NTD pseudovirus mutants, (B) RBD pseudovirus mutants, and (C) S1/S2 pseudovirus mutants. Particles pseudo-typed with the indicated S proteins were pre-incubated for one hour at 37 °C with plasma dilutions from health care workers vaccinated with BNT162b2 XBB.1.5 adapted vaccine. The numbers of biological replicates corresponding to individual plasma samples are shown on top and were assessed in technical duplicates. Pseudo-virus neutralization titer 50 (PVNT50) was calculated using the least squares fit using a variable slope, using a four-parameter nonlinear regression model and values were plotted as geometric mean. Geometric mean standard deviation bars are shown in black. The lower limit of confidence (LLOC) was set at a PVNT50 of 50. Non responders are defined as individuals below 60 (dashed line). All PVNT50 below 10 are set at 10 for visualization purposes. The assay was performed with negative controls to assess the virus input of each used pseudo-virus in the absence of plasma antibodies. Geometric means and fold change neutralization over $BA.2_{pp}$ are shown on top.

### Supplementary figure 3

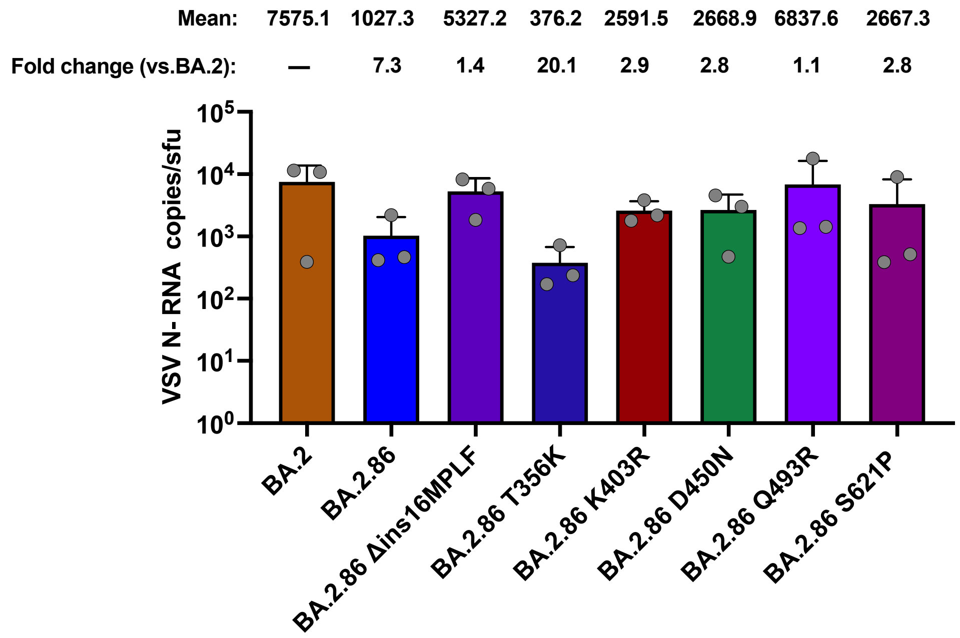


**Figure S3: ddPCR results of SARS CoV-2 VSV pseudovirus N- RNA copy number of pseudovirus stocks**

Viral RNA was extracted from three independent stocks from each shown pseudovirus. Supernatants were pretreated with nuclease to remove unpackaged RNA. VSV N-protein RNA genome copy number was assessed by ddPCR. Results are represented as total VSV N- gene copy number per single infectious unit.
